# Incidence, predictors and cognitive dysfunction in post-traumatic epilepsy following a traumatic brain injury: A study protocol

**DOI:** 10.1101/2022.11.10.22282171

**Authors:** Irma Wati Ngadimon, Mohd. Farooq Shaikh, Devi Mohan, Ching Soong Khoo, Wing Loong Cheong, Angel Aledo-Serrano

## Abstract

Approximately 69 million people worldwide are annually affected by traumatic brain injury (TBI). In Malaysia, the traumatic injury was the leading cause of hospital admission and death, accounting for one in three emergency visits. Among the most recognised complication of TBI is post-traumatic epilepsy (PTE), which is an essential contributor to morbidity and mortality. However, there is a lack of local epidemiological data on PTE in Malaysia. This study aims to describe the incidence and predictors of PTE among TBI patients admitted to a tertiary healthcare centre in Kuala Lumpur, Malaysia. We hypothesised that increases in age, race, and severity of brain injury are among the main potential predictors of PTE. It will also provide evidence that patients with epilepsy following TBI are associated with significant impairment in cognitive performance than TBI patients without epilepsy. An analysis of a two years retrospective cohort will be employed, of which adults with a history of admission for TBI in 2019 and 2020 will be contacted, and the development of epilepsy will be ascertained using a validated tool and confirmed by our neurologists during visits. The patients will then be grouped into two, with PTE and without PTE, and assessed their cognitive performance by clinical psychologists. Given that the management of TBI and PTE patients involves a multidisciplinary team, the findings might be significant to many healthcare providers in determining policy and strategise a better treatment.

## Introduction

At least 10 million people sustain head trauma every year [2,3], making traumatic brain injury (TBI) one of the leading global causes of death and disease burden [1]. One of TBI’s major complications but poorly understood sequelae is post-traumatic epilepsy (PTE) [4–6]. PTE is a significant form of symptomatic epilepsies resulting from various brain insults [6]. In the general population, it causes 10–20% of symptomatic epilepsy and 5% of all epilepsy [7].

PTE poses significant health and socio-economic problem worldwide [8]. People with epilepsy are three times more likely to die prematurely than the general population, with the absolute number of deaths increasing by 39% [1]. Post-traumatic epileptic seizures are distinguished from non-epileptic post-traumatic seizures (PTS) by their etiology and timing after the trauma. Late seizures, occurring more than a week after the initial event, characterise PTE. [9]. The time between a TBI and the onset of the first seizure varies greatly [10]. In about 80% of cases, the first seizure following PTE occurs within the first year, and more than 90% of cases occur by the end of the second year [11]. Eighty-two percent of patients who experienced a late-onset seizure (more than a week after injury) also experienced a second seizure within the following two years [8]. Although a seizure during the first week after injury (early PTS) is linked to a higher incidence of late PTS (after the first week of injury), having a late PTS is correlated with an even higher risk of seizure recurrence [12–14]. The likelihood of seizures following a TBI has been predicted mathematically by several authors, but their clinical applicability for the majority of brain-injured people has not been tested [13]. Researchers have tried to pin down the true prevalence of PTE for decades, and their findings have yielded a wide range of reported numbers, from 1.3% to 53.3% [15–17]. This variation may be attributable to differences in the severity of the initial TBI. The cumulative incidence of PTE was reported to be 4.4 per 100 people with mild TBI, 7.6% per 100 people with moderate TBI, and 13.6 per 100 people with severe TBI in a three-year population-based study conducted in the United States [4].

In most TBI cases, cognitive difficulties are the most prominent impairments [18]. However, little is known about how PTE can exaggerate neuropsychological deficits. Preclinical TBI studies provided scientific proof that PTE may further deteriorate cognitive performance. This comorbidity is also reported in other secondary epilepsies, such as post-stroke epilepsy [19]. Cognitive performance includes motor function, memory, intelligence, attention or concentration, processing speed, language, and visual-spatial abilities [20]. Researchers found that mice induced with TBI followed by electroconvulsive shock seizures performed worse in the Barnes maze than mice who underwent TBI alone [21]. These findings on the “double hit” mice were related to increased glial activation.

Meanwhile, in a study by Shultz and colleagues [22] comparing rats that did or did not develop PTE at six months post-injury, no significant differences between the groups were observed in Morris water maze (MWM) performance. The few preclinical studies that evaluated the impacts of PTE on cognitive functions have yielded inconclusive results. In some studies, temporal complexities may contribute to the failure to distinguish between PTE versus non-PTE groups of post-TBI rats. One previous study found an alarming statistic: half of the people with epilepsy exhibit cognitive dysfunctions in multiple domains, including learning, memory, attention, and executive functioning [23]. Memory loss, however, is far more common [24]. As a result, it is prudent to postulate that epilepsy resulting from TBI adds deficits to an individual with TBI, thereby worsening neuropsychological performance.

This study involves two parts. Part I of the study is to determine the etiology of PTE in TBI individuals to quantify the incidence rate and stratify risks for developing PTE. Part II of the study examines the cognitive comorbidity of PTE in a diverse multi-ethnic population. Previously, several authors [5,6,13,25,26] had analysed predictors for PTE in a large sample of TBI people. Unfortunately, the geographical limitation prevents the broad generalisation of the results of these investigations. To the best of our knowledge, this study represents the first investigation in Malaysia with multi-ethnicity and various demographical backgrounds who were retrospectively reviewed for up to 2 years after brain injury. The results of this study are intended to add to the emerging body of knowledge for both professionals and the general public. Individuals with TBI and PTE face unique challenges that healthcare providers will consider. Due to this, better services and strategies to enhance these people’s lives could be created. Additionally, people with PTE and their families may be better prepared to deal with upcoming issues such as memory deficit, driving limits, intelligence, and the effect on caregivers, family members, and significant others. Furthermore, reducing the burden of comorbidities linked with decreased cognitive abilities and PTE would substantially impact the individual, society, and the economy.

## Significance of the study

Despite a wealth of epidemiological data, there are still many questions regarding the epidemiology and determinants of outcomes of TBI patients with no definitive answers and no robust evidence base to guide their management. There is a relative lack of data on PTE in Malaysia. As a consequence, it has become an underappreciated condition. Patients with similar injuries may receive very different opinions about their likely prognosis and the best treatment, depending on their unit and the surgeon who treats them. Once a risk has been unequivocally associated with PTE, a logical final step is targeting those individuals with specific predictors in advocating a policy that aims to strategise the pharmacological or non-pharmacological treatment toward preventing epilepsy and its cognitive comorbidity. The findings can be used to plan and evaluate strategies to prevent further deterioration and guide the management of patients whom PTE has already developed, thereby helping to lighten the burden of ill health upon the individual and the community.

## Aim and objectives

### Study aim

This project aims to describe the incidence and predictors for PTE among TBI in a tertiary care center in Malaysia and generate evidence on the cognitive burden of PTE.

### Study objectives

a. To estimate the incidence of PTE among TBI patients attending Universiti Kebangsaan Malaysia Medical Centre (UKMMC), Kuala Lumpur, between 2019 to 2020.
b. To identify potential predictors of PTE among TBI patients in UKMMC, Kuala Lumpur.
c. To determine the association between PTE and cognitive impairment in the study population

## Research Question

i. What is the incidence of PTE among the TBI patients attending UKMMC, Kuala Lumpur, between 2019 to 2020?
ii. What are the potential predictors of PTE among the TBI population in UKMMC, Kuala Lumpur?
iii. Does PTE correlate with cognitive impairment in TBI patients?

## Materials and method

### Study design and setting

This retrospective cohort study will take place in UKMMC, Kuala Lumpur, from March 2022 until March 2024. All TBI patients admitted from 2019 to 2020 to UKMMC that meet the inclusion criteria will be retrieved from the hospital ward census.

### Inclusion criteria

Inclusion criteria are (a) above 18 years old, (b) patient or guardian able to understand and speak English/Malay, and (c) with a history of admission to the Neurosurgery ward in UKMMC for traumatic brain injury caused between 2019 to 2020.

### Exclusion criteria

Patients are excluded if (a) the patient or guardian does not understand Malay/English, (b). declines to participate, (c) individuals with pre-existing epilepsy before TBI, (d) brain disorder that may cause epilepsy, e.g., brain tumor, stroke, meningitis, dementia.

### Sample and sampling

We will approach All TBI patients meeting the inclusion criteria retrieved from the ward census.

#### a) Study Part I: Determine the incidence and predictors of PTE

##### Sample size calculation

The reported incidence of PTE range from about 1% to 18.5% in year 1 [8,27–30]. Hence for a cumulative incidence of 18.5%, the sample size for 95% CI is 232 with an alpha error of 5% (https://www.openepi.com/SampleSize/SSPropor.htm). Assuming a drop out of 10%, thus 232+ 23= 255, rounded off to 260. Therefore, the TBI subjects to be recruited are 260.

##### Data collection process

The study flowchart is depicted in Fig 1. A TBI database will be formed based on the list. The patient’s medical records in UKMMC will be retrieved from the medical record unit to find related demographic and clinical information, which are:

**Figure.**
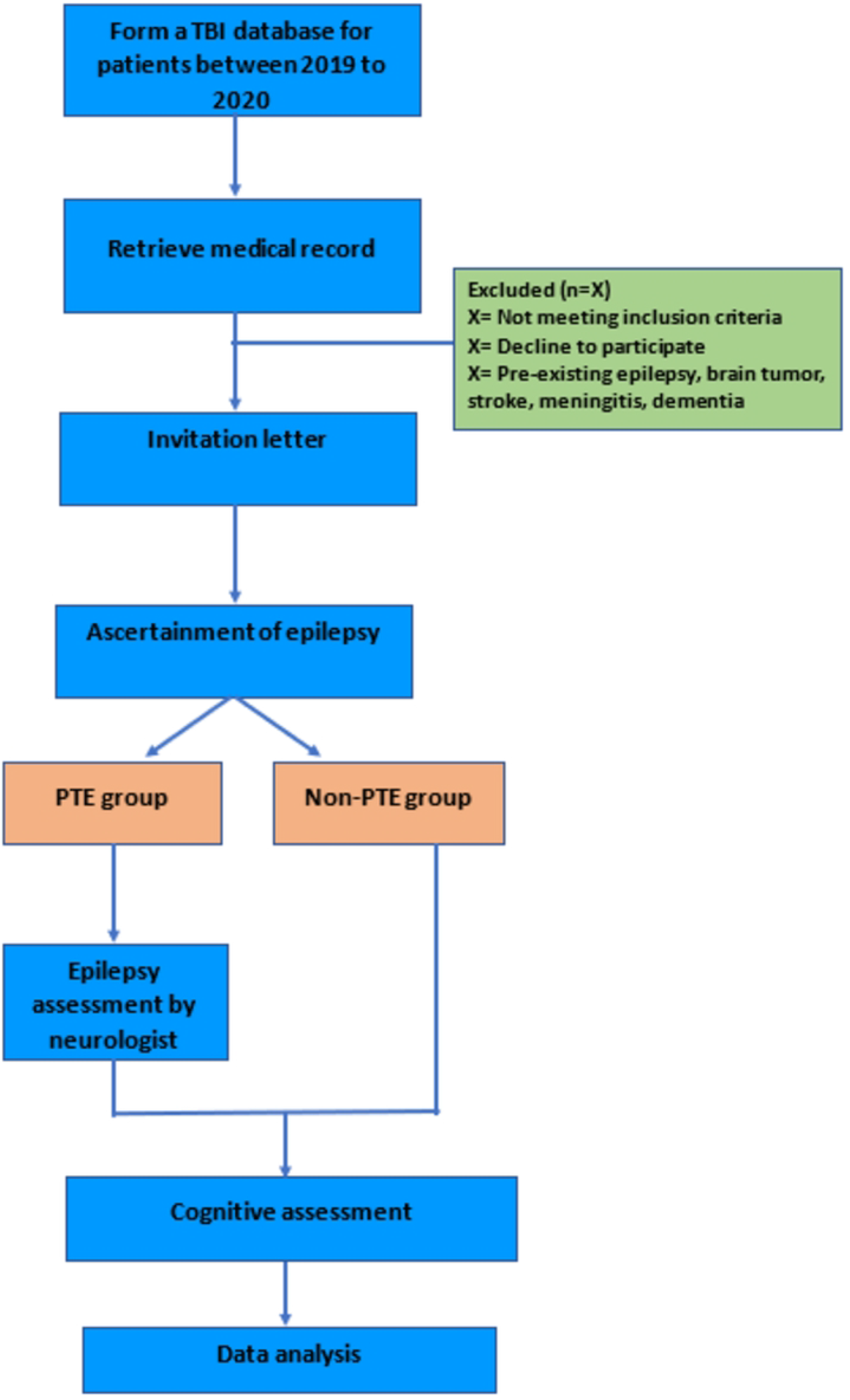

- Demographic and basal variables: Age, gender, ethnicity, comorbid illness, and medical history focus on neurological conditions.
- TBI characteristics during admission: Glasgow Coma Scale (GCS), loss of consciousness, vomiting, post-traumatic seizure (PTS), cranial surgery, and neuroimaging (looking for different aspects, such as subdural hematoma (SDH), subarachnoid hemorrhage (SAH), extradural hemorrhage (EDH), Intraparenchymal hemorrhage (IPH), midline shift, hydrocephalus, cerebral edema, and skull fracture).
- Seizure and epilepsy evaluation:

Seizures occurrence in-ward (if any), type, duration of seizure episode, EEG monitoring for evidence of epileptogenesis, epilepsy diagnosis, and anti-seizure medication.

An invitation letter that mentions the researcher’s background, the study’s aims, and the design will be sent to the patients’ home addresses. Next, patients will be contacted by telephone to ascertain their epilepsy using a validated instrument adapted from a study by Ottman et al. [31] and the Malay version by Fong et al. [32]. Permission to use the original screening instruments has been obtained from both authors.

We made some modifications to the scoring system as below. We classified the response as positive having epilepsy following TBI if the subject answered as:

i. Q1 and Q3d: “No” or “don’t know” and
ii. Any Q2, Q3a, b, c, e, f, g: “Yes” or “possible.”

The following questions were also asked of patients with positive symptoms. Responses were recorded in a subject data sheet (APPENDIX III).

i. Before the TBI, did the patient experience seizures or epilepsy?
ii. Have the most recent seizure(s) occurred more than seven days after the TBI?
iii. Has the patient been diagnosed with epilepsy or a seizure disorder since the TBI diagnosis? If so, when was the date of diagnosis, and which clinician/hospital made the diagnosis?
iv. Any anti-seizure /anti-epileptic medication received?

All screened cases will be discussed with a consultant neurologist. Patients who declared having symptoms of PTE but have not sought any treatment determined by a record of follow-up in UKMMC will be directed to a neurologist for clinical observation and further treatment. Electroencephalography (EEG) will be used as an adjunct diagnostic tool. Then, patients who consented to cognitive assessment will be grouped into two, with PTE and without PTE. The cognitive assessment will be performed by our clinical psychologist and trained researcher at Neurology Clinic UKMMC or by home visit.

##### Consent taking

The consent-taking process will be done verbally at the beginning of the phone call (APPENDIX IV). First, the researcher will introduce herself and explain the purpose of the phone call. Then, the researcher will play a pre-recorded Patient Information Sheet (PIS) script to ensure transparency (APPENDIX V). The patient/ guardian will be given sufficient time to think and ask questions afterward. Next, verbal consent from the patient or guardian will be taken before the interview, and the process will be recorded. The phone call will be discontinued if no permission is given. Suppose the patient is not fit to be interviewed (e.g., hearing impairment or cognitively not fit to answer); in that case, the patient’s guardian will be contacted.

#### b) Study Part II: Cognitive assessments

##### Sample size calculation

The next part of the study is the sub-analysis of cognitive performance. The ratio of individuals from Part I in the PTE group to individuals in the non-PTE group (as the control group) to be recruited is 1:3. The number of PTE patients depends upon the incidence within the sample of 260. For example, the incidence of PTE is 18.5% which means 48, so 48 subjects in the PTE group vs. 144 subjects in the non-PTE group. The control group will be age-matched and gender-matched with the study group.

##### Data collection process

The following is the cognitive assessments and anxiety/depression test involved:

i. Addenbrooke’s Cognitive Examination-III (ACE-III) test [33] with both English and Malay versions [34].
ii. Wechsler Adult Intelligence Scale (WAIS-IV) subtest: symbol search and coding [35].
iii. Comprehensive Trail Making Test (CTMT) [36,37].
iv. Depression, Anxiety and Stress Scale (DASS 21) [38,39].

##### Data analysis

Statistical analysis will be performed using SPSS 28.0. Chi-square test or Fisher exact test will be conducted to compare categorical data, independent sample t-test for continuous variables, and ANOVA with repeated measures to study between subjects’ factor group, within subjects’ factor time, and post-doc tests with Bonferroni correction method. Pearson correlations and logistic regression analysis will determine if the variables are significantly related. The cognitive score difference between the two groups will be analysed using SPSS.

##### Ethics approval

This study has been approved by the Research Ethics Committee, The National University of Malaysia (UKM PPI/111/8/JEP-2022-072).

##### Status and timeline of study

The TBI patients’ database has been formed, and participant recruitment is in progress. The study is expected to be completed by June 2023. The data analysis will be performed by July 2023, and the research article will be completed by September 2023.

## Discussion

This study will benefit a multidisciplinary team involved in managing TBI patients, like critical care physicians, neurosurgeons, neurologist rehabilitation specialists, and clinical psychologists. The results will help clinicians to understand the best way to treat TBI and PTE cases and serve as a source of information to support care decisions in designing therapies that would improve the quality of life of TBI patients. There are several limitations of this study. It is a single-center study; thus, the sample may not represent all geographic regions of Malaysia. Nonetheless, to our knowledge, our study constitutes the first analysis of PTE incidence in Malaysia. As with the other retrospective study designs, the results will rely on whatever information is available in the medical record during data collection. Thus, there might be unrecorded or unavailable information. To deal with it, we will try to capture important details such as the patients’ TBI history, education level, and any signs and symptoms of seizure/epilepsy during the phone interview session.

We anticipate that patients willing to come for cognitive assessment might be cognitively well, thus leading to the skewness of results. We offer a home visit for individuals with ambulatory problems to minimise bias.

Another challenge is that the contacts provided may no longer be in service. The previous study shows that some may not want to answer calls due to business or worry about answering unknown phone numbers [40]. To reduce this problem, we will send a letter to the individual’s home address to mention our study purpose and action plan to contact them. In the letter, we will provide the team telephone numbers and a Q.R. code linked to Whatsapp so they can propose their preferred date and time for a phone interview. A token of RM20 will also be provided to cover the petrol cost of patients participating in the cognitive assessments.

## Data Availability

No datasets were generated or analysed during the current study. All relevant data from this study will be made available upon study completion.

## Author’s contributions

IWN: Data curation, Writing, Investigation, Formal Analysis

MFS: Conceptualization, Project Administration, Funding Acquisition

DM: Conceptualisation, Formal Analysis, Supervision

CSK: Investigation, Supervision

WLC: Validation, Supervision

AAS: Validation, Supervision

## Acknowledgements

This work forms part of the first author’s Ph.D. IWN.

## Supporting information

Appendix I: Ascertainment of epilepsy by Ottomen et al., 2010

Appendix II: Ascertainment of epilepsy by Fong et al., 2019

Appendix III: Subject data sheet

Appendix IV: Telephone script for verbal consent and recruitment

Appendix V: Patient Information Sheet (PIS)

## Notes

### Competing Interest Statement

The authors have declared no competing interest.

### Funding Statement

This project is part of bigger study supported by Jeffrey Cheah School of Medicine and Health Science Strategic Grant 2021, Monash University Malaysia. Mohd. Farooq Shaikh is the principal investigator. Website: https://www.monash.edu.my/jcsmhs The funders had and will not have a role in study design, data collection and analysis, decision to publish, or preparation of the manuscript.

